# Long-Term Impact of Digital Media on Brain Development in Children

**DOI:** 10.1101/2022.07.01.22277142

**Authors:** Samson Nivins, Bruno Sauce, Magnus Liebherr, Nicholas Judd, Torkel Klingberg

**Author notes:** Corresponding authors: Dr. Samson Nivins, Department of Neuroscience (Neuro),C4, Karolinska Institutet, Sweden, Prof. Torkel Klingberg, Department of Neuroscience, Karolinska Institutet, Sweden.

## Abstract

Digital media (DM) takes an increasingly large part of children’s time, yet the long-term effect on brain development remains unclear. We investigated how individual effects of DM use (i.e., using social media, playing video games, or watching television/videos) on the development of the cortex (i.e., global cortical surface area), striatum, and cerebellum in children over four years, accounting for both socioeconomic status and genetic predisposition. We used a prospective, multicentre, longitudinal cohort of children from the Adolescent Brain and Cognitive Development Study, aged 9.9 years when entering the study, and who were followed for four years. Annually, children reported their DM usage through the Youth Screen Time Survey and underwent brain magnetic resonance imaging scans every two years. Quadratic-mixed effect modelling was used to investigate the relationship between individual DM usage and brain development. We found that individual DM usage did not alter the development of cortex or striatum volumes. However, high social media usage was associated with a statistically significant change in the developmental trajectory of cerebellum volumes, and the accumulated effect of high-vs-low social media users on cerebellum volumes over four years was only β= −0.03, which was considered insignificant. Nevertheless, the developmental trend for heavy social media users was accelerated at later time points. This calls for further studies and longer follow-ups on the impact of social media on brain development.

## Introduction

Children are increasingly engaged with digital media (DM) more than ever before. For example, in the U.S., children aged 8-12 years, on average, spend 4 hours and 44 minutes daily on DM for entertainment purposes,^1^ in addition to its use during school and homework. This rise in usage has sparked concerns among parents, caregivers, and policymakers regarding its potential adverse effects on the developing brains of children. However, research in this domain remains inconclusive and somewhat contradictory.

Concerning the DM’s impact on cognitive outcomes, prior studies have reported both beneficial and detrimental associations.^2–5^ Similarly, a recent review on brain development simply noted that DM’s effects can be both positive and negative.^6^ This inconsistency in findings can be attributed to several factors. First, the general term ‘digital media’ encompasses a wide range of activities, each potentially influencing development in distinct ways or even exerting contrasting effects. Therefore, it is crucial to differentiate between various digital activities, such as playing video games, watching television/videos, and using social media. Second, the age of the participants is a significant factor. For example, research by Orben et al. in 2022 showed that social media use could negatively affect psychological well-being during particular developmental stages, with these stages occurring at different times for boys and girls.^7^ In another study, Soares et al. found that boys who spent more time watching television or playing video games at 11 years old, and more time using computers at 11 and 15 years old, showed improved working memory performance at 22 years old.^8^ However, this association was not observed in girls. Third, and perhaps most critical, is the conflation of evidence from cross-sectional and longitudinal studies in reviews. Cross-sectional studies can identify correlations but cannot establish causality. Whereas longitudinal studies may even yield opposite results. For example, a longitudinal study using structural equation modeling has found a negative correlation between time spent playing video games and intelligence.^4^ However, when controlling for baseline cognition and other background variables, the longitudinal analysis revealed that playing video games positively influenced changes in intelligence (β=0.17). The initial negative correlation between video gaming and cognitive performance was interpreted as resulting from self-selection.

Longitudinal research on the effect of DM and brain development in children remains limited. A series of studies on a cohort of Japanese children observed that watching television increased grey matter volume in frontal areas,^9^ playing video games increased mean diffusivity in the white matter,^10^ and internet usage decreased grey matter volume in extensive brain regions.^11^ Although informative, this is a single cohort of less than 300 individuals, which varied widely in age, between ages 6 to 18. Brain development during this period is nonlinear, which was not accounted for in the statistical modeling. In 2023, Miller et al. assessed the impact of DM on functional connectivity over two years in a cohort of over 4000 children.^12^ They reported no effects exceeding a size of 0.2, the predetermined threshold for significance.

The ongoing debate over what constitutes a meaningful effect size continues without consensus in psychology and neuroscience. This issue is particularly relevant in large-scale studies like the Adolescent Brain Cognitive Development (ABCD) study, where statistical significance may not equate to a meaningful effect for the individual.^13^ The traditional criteria by Cohen, which categorizes effect sizes of 0.2 as small and 0.5 as medium, were arbitrary from the outset, with Cohen himself acknowledging the lack of solid evidence for these benchmarks.^14^ Funder and Ozer propose that effect sizes must be contextualized, and propose as guidelines that an r-value of 0.05 indicates a very small effect and 0.1 a small effect. The frequency of an event may also be crucial, as repeated events can accumulate effects over time, according to Abelson.^15^

Furthermore, when interpreting effect sizes, it is essential to consider additional factors. Even a small effect can have significant implications if it influences various aspects of an individual’s life or interacts positively with other variables. Habituation or counteractive responses might mitigate an effect’s impact.^16^ In our analysis, we regard an annual effect size of 0.05 as meaningful. This threshold is deemed appropriate, considering the cumulative influence of DM and the potential for an effect on a general ability like attention to significantly impact schooling and everyday life.

Our study aimed to investigate the individual effects of DM usage on structural brain development in children aged 9.9 years at baseline (T_0_) over four years, adjusted for age, sex, SES, scanner sites, and genetic predisposition. We selected global cortical surface area (CSA) as the main outcome measure since previous studies have shown a strong relationship between global CSA and intelligence across different age groups.^17–22^ Moreover, we used cortical surface area rather than cortical thickness because studies have consistently reported an association of environmental variables, such as SES, on cortical surface area rather than on cortical thickness.^23^

We further investigated the individual brain structures, i.e., the volumes of the striatum and cerebellum, which have been implicated in cross-sectional studies of DM usage.^24,25^ In general, global CSA tends to increase during this period of childhood a part of normal development with a peak age at 11 years of age.^26^ Building upon our prior research findings,^4^ we hypothesized that DM usage, particularly playing video games, would be associated with an increase in global CSA. Since DM usage differs between sexes,^27^ we will study the effect of sex on these relationships.

## Results

### Baseline characteristics

Of the 11,875 children from the ABCD study cohort, 6469 children (age, mean (SD) = 9.9 (0.6) years; boys, n (%) = 3369 (52.1%)) fulfilled our inclusion criteria and were included at the T_0_ visit (i.e., 9-11 years of age). Of these, 4610 children (age = 12.0 (0.7) years; boys = 2487 (53.9%)) were included at the T_2_ (i.e., two years later) visit, and 1697 children (age = 13.4 (0.6) years; boys = 949 (55.9%)) were included at the T_4_ (i.e., four years later) visit. 1462 children had usable data for all three-time points.

The estimated time spent by these children on DM types at T_0_ was 0.5 hours/day for using social media, 0.9 hours/day for playing video games, and 2.1 hours/day for watching television/videos **(Table 1)**.

**Table 1.**
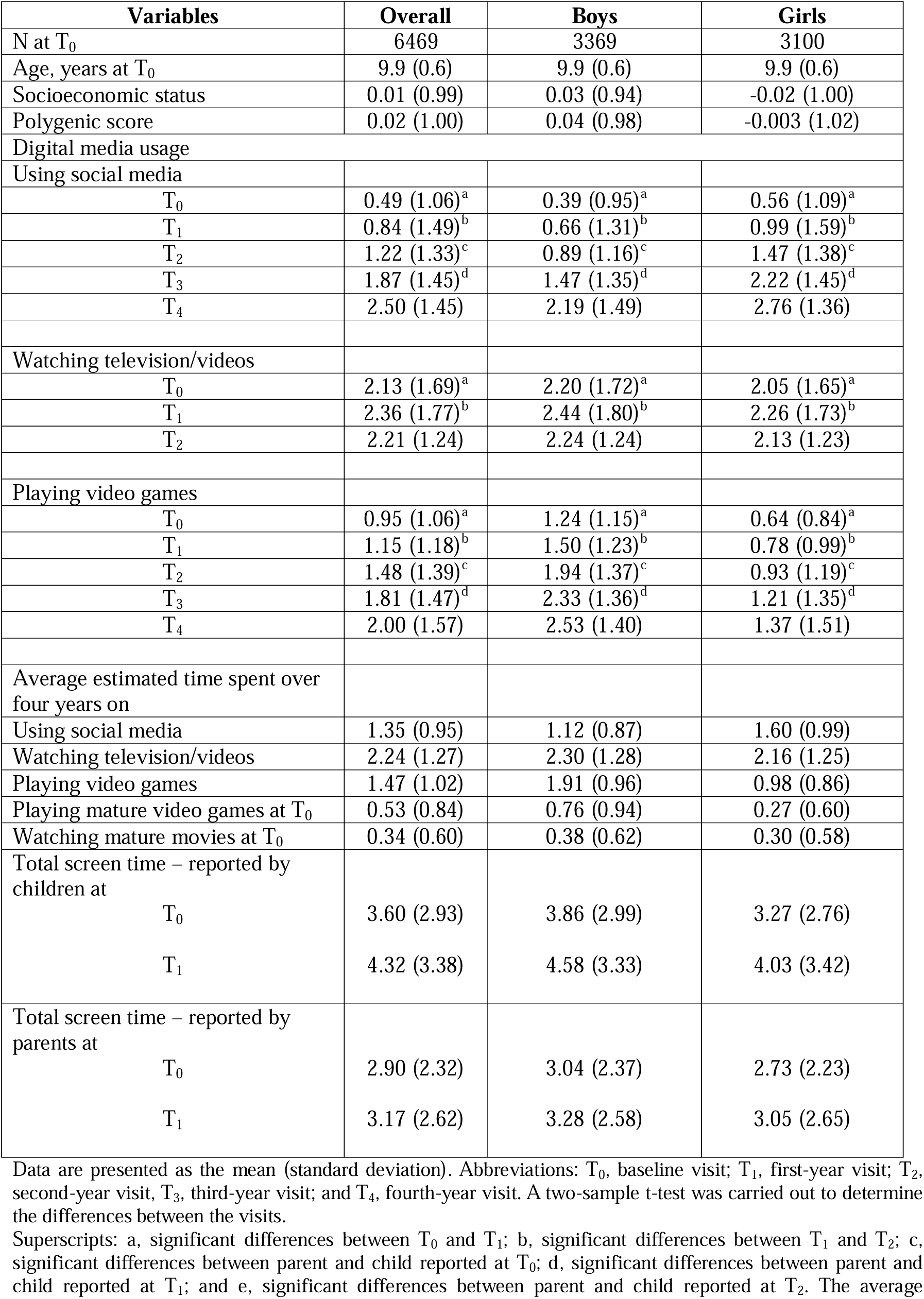
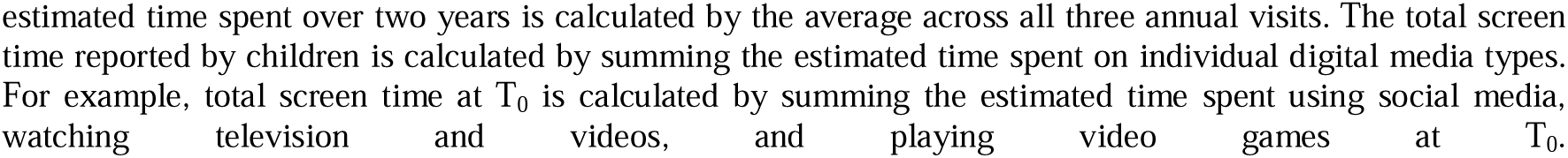
Descriptive characteristics of the sample.

| Variables | Overall | Boys | Girls |
| --- | --- | --- | --- |
| N at T<sub>0</sub> | 6469 | 3369 | 3100 |
| Age, years at T<sub>0</sub> | 9.9 (0.6) | 9.9 (0.6) | 9.9 (0.6) |
| Socioeconomic status | 0.01 (0.99) | 0.03 (0.94) | -0.02 (1.00) |
| Polygenic score | 0.02 (1.00) | 0.04 (0.98) | -0.003 (1.02) |
| Digital media usage |  |  |  |
| Using social media |  |  |  |
| T<sub>0</sub> | 0.49 (1.06)<sup>a</sup> | 0.39 (0.95)<sup>a</sup> | 0.56 (1.09)<sup>a</sup> |
| T<sub>1</sub> | 0.84 (1.49)<sup>b</sup> | 0.66 (1.31)<sup>b</sup> | 0.99 (1.59)<sup>b</sup> |
| T<sub>2</sub> | 1.22 (1.33)<sup>c</sup> | 0.89 (1.16)<sup>c</sup> | 1.47 (1.38)<sup>c</sup> |
| T<sub>3</sub> | 1.87 (1.45)<sup>d</sup> | 1.47 (1.35)<sup>d</sup> | 2.22 (1.45)<sup>d</sup> |
| T<sub>4</sub> | 2.50 (1.45) | 2.19 (1.49) | 2.76 (1.36) |
| Watching television/videos |  |  |  |
| T<sub>0</sub> | 2.13 (1.69)<sup>a</sup> | 2.20 (1.72)<sup>a</sup> | 2.05 (1.65)<sup>a</sup> |
| T<sub>1</sub> | 2.36 (1.77)<sup>b</sup> | 2.44 (1.80)<sup>b</sup> | 2.26 (1.73)<sup>b</sup> |
| T<sub>2</sub> | 2.21 (1.24) | 2.24 (1.24) | 2.13 (1.23) |
| Playing video games |  |  |  |
| T<sub>0</sub> | 0.95 (1.06)<sup>a</sup> | 1.24 (1.15)<sup>a</sup> | 0.64 (0.84)<sup>a</sup> |
| T<sub>1</sub> | 1.15 (1.18)<sup>b</sup> | 1.50 (1.23)<sup>b</sup> | 0.78 (0.99)<sup>b</sup> |
| T<sub>2</sub> | 1.48 (1.39)<sup>c</sup> | 1.94 (1.37)<sup>c</sup> | 0.93 (1.19)<sup>c</sup> |
| T<sub>3</sub> | 1.81 (1.47)<sup>d</sup> | 2.33 (1.36)<sup>d</sup> | 1.21 (1.35)<sup>d</sup> |
| T<sub>4</sub> | 2.00 (1.57) | 2.53 (1.40) | 1.37 (1.51) |
| Average estimated time spent over four years on |  |  |  |
| Using social media | 1.35 (0.95) | 1.12 (0.87) | 1.60 (0.99) |
| Watching television/videos | 2.24 (1.27) | 2.30 (1.28) | 2.16 (1.25) |
| Playing video games | 1.47 (1.02) | 1.91 (0.96) | 0.98 (0.86) |
| Playing mature video games at T<sub>0</sub> | 0.53 (0.84) | 0.76 (0.94) | 0.27 (0.60) |
| Watching mature movies at T<sub>0</sub> | 0.34 (0.60) | 0.38 (0.62) | 0.30 (0.58) |
| Total screen time – reported by children at |  |  |  |
| T<sub>0</sub> | 3.60 (2.93) | 3.86 (2.99) | 3.27 (2.76) |
| T<sub>1</sub> | 4.32 (3.38) | 4.58 (3.33) | 4.03 (3.42) |
| Total screen time – reported by parents at |  |  |  |
| T<sub>0</sub> | 2.90 (2.32) | 3.04 (2.37) | 2.73 (2.23) |
| T<sub>1</sub> | 3.17 (2.62) | 3.28 (2.58) | 3.05 (2.65) |

Data are presented as the mean (standard deviation). Abbreviations: T<sub>0</sub>, baseline visit; T<sub>1</sub>, first-year visit; T<sub>2</sub>, second-year visit; T<sub>3</sub>, third-year visit; and T<sub>4</sub>, fourth-year visit. A two-sample t-test was carried out to determine the differences between the visits.
Superscripts: a, significant differences between T<sub>0</sub> and T<sub>1</sub>; b, significant differences between T<sub>1</sub> and T<sub>2</sub>; c, significant differences between parent and child reported at T<sub>0</sub>; d, significant differences between parent and child reported at T<sub>1</sub>; and e, significant differences between parent and child reported at T<sub>2</sub>. The average
estimated time spent over two years is calculated by the average across all three annual visits. The total screen time reported by children is calculated by summing the estimated time spent on individual digital media types. For example, total screen time at $T_0$ is calculated by summing the estimated time spent using social media, watching television and videos, and playing video games at $T_0$ .

Compared to the T_0_ visit, the estimated time spent using DM types significantly increased over four years in the overall cohort and in boys and girls (**Table 1)**. During the four years of the follow-up period (i.e., across all four annual visits), children, on average, spent 1.4 hours/day using social media, 1.5 hours/day playing video games, and 2.2 hours/day watching television/videos. Moreover, during this period, boys spent more time playing video games or watching television/videos, whereas girls spent more time using social media or watching television/videos.

As expected, parents reported less total screen time use per day in children compared to child reports across two visits **(Table 1)**.

### Normal brain developmental trajectory

Overall development followed an inverted U-shaped developmental trend for global CSA, striatum and cerebellum volumes between mid-childhood and early adolescence, i.e., age-related increase during mid-childhood and subsequently decrease during early adolescence. According to the fitted model, the global CSA, striatum, and cerebellum volumes peaked at 10.6, 10.9, and 15.4 years, respectively **(Figure 1a)**.

**Figure 1.**
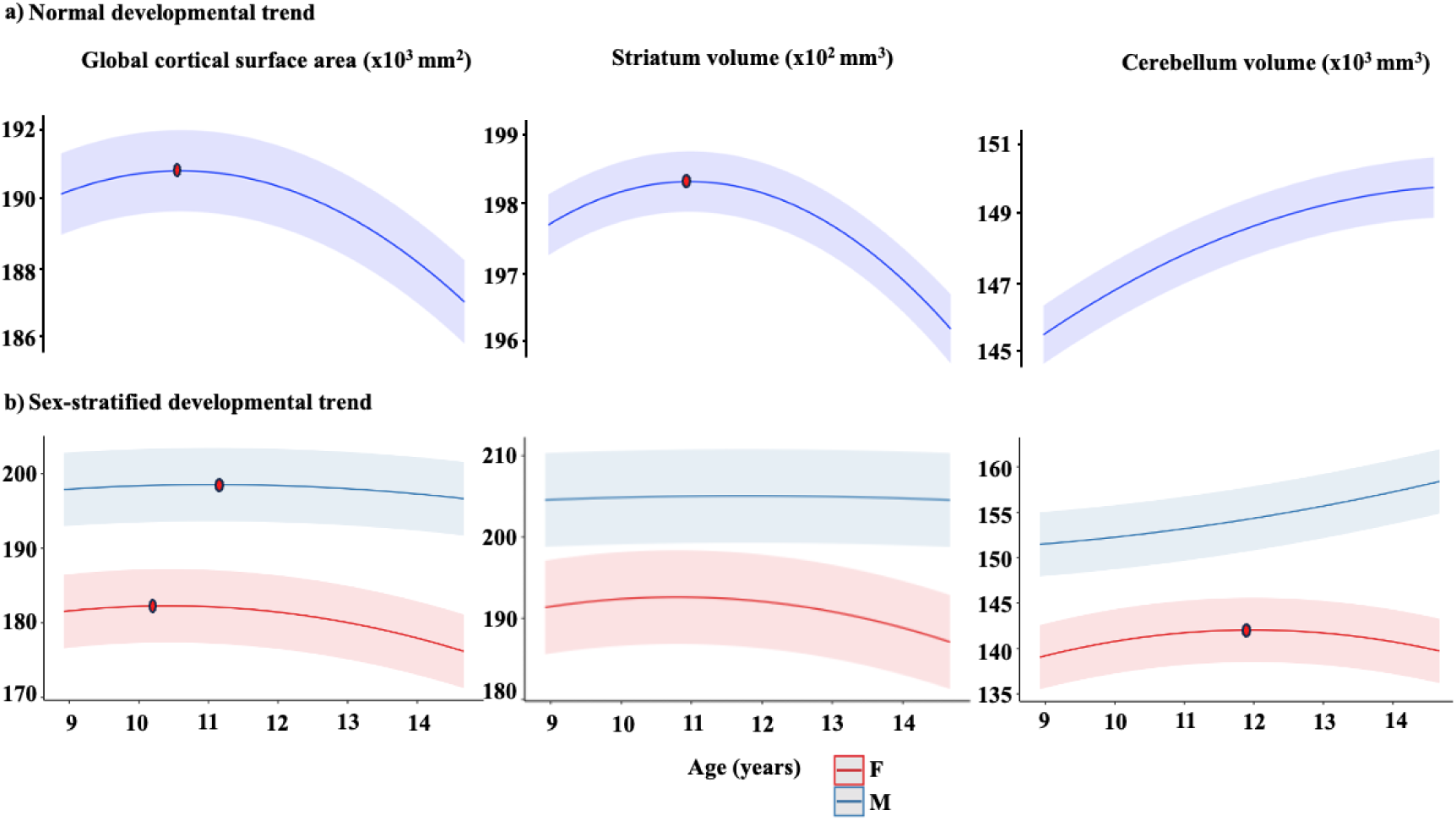
**a,** Plots representing the quadratic effects of age (years) predicting global cortical surface area, striatum and cerebellum volumes in the overall cohort, adjusted for sex, socioeconomic status, sex, polygenic scores for cognitive performance, and 20 principal components, and the grey shade around the regression lines corresponds to a 95% confidence interval of the intercept; **b**, sex-stratified developmental trend adjusted for the same covariates as mentioned above. The dots represent the peak age, estimated by the first derivative. The y-axis represents brain structures.

### Sex effect on brain development

To determine whether the trajectory differed between the sexes, we added interaction terms (age * sex; age^2^ * sex) to the pre-existing model. There was a significant interaction for sex with global CSA and cerebellum volumes, but not for striatum **(eTable 2)**.

Overall, boys had larger global CSA and cerebellum volumes than girls **(eTable 3; Figure 1b)**, but the peak was much earlier in girls (global CSA = 10.4 years and cerebellum = 11.9 years) than in boys (global CSA = 11.1 years). Cerebellum volumes increased over this age range for boys.

### SES and cogPGS

Overall, we observed a positive association between SES and global CSA (β = 0.08 to 0.11; p < 0.001) (**Table 2)**. For illustration purposes, we additionally categorized SES into quartiles and plotted global CSA development across them **(Figure 2)**. Similarly, we categorized SES into three quartiles and studied the trend for cerebellum volumes (**Table 2).** Overall, children from lower levels of SES had lower global CSA and cerebellum volumes compared with their developing peers and had earlier maturation **(Figure 3)**.

**Figure 2.**
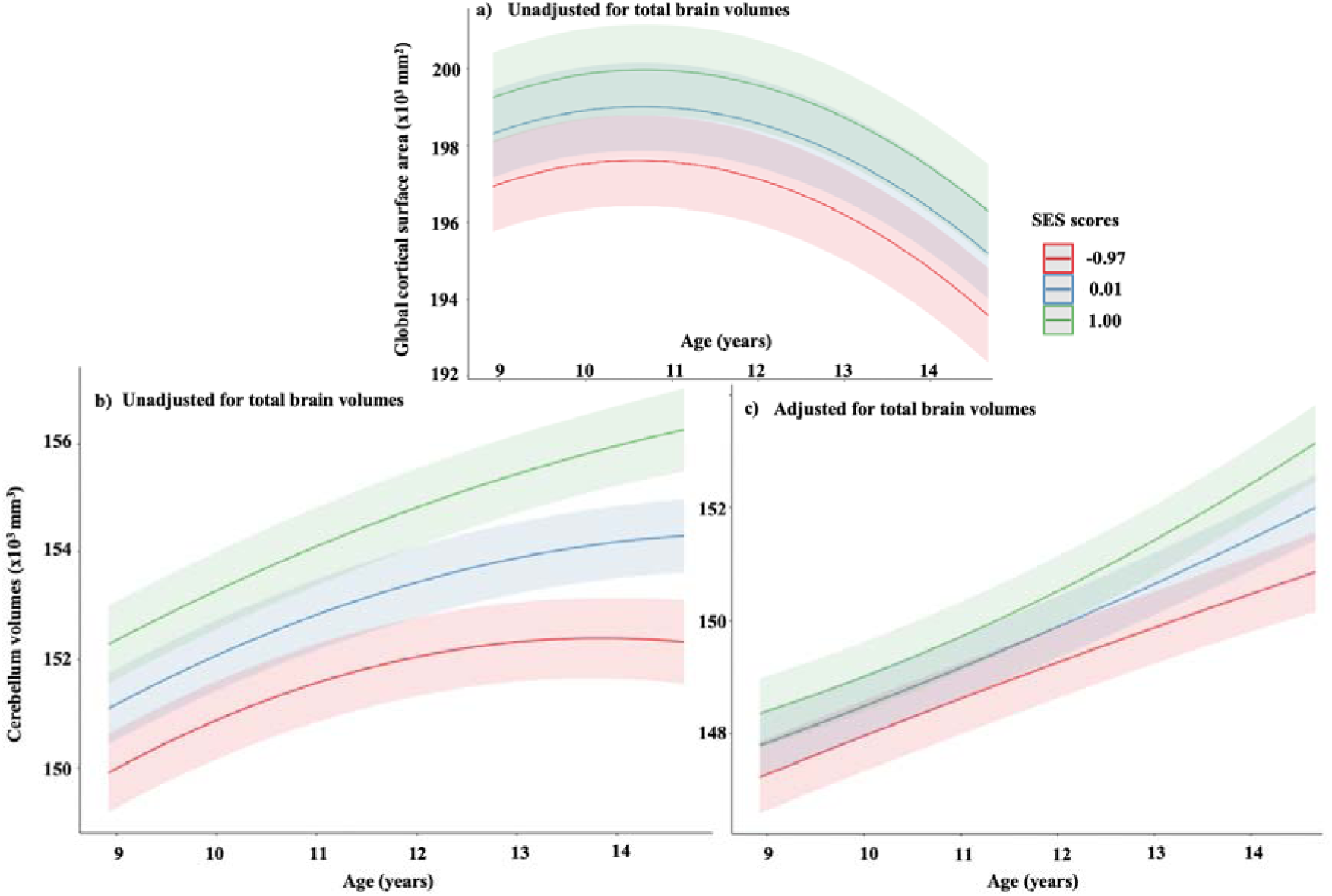
Developmental trajectories and socioeconomic status, **(a)** global cortical surface area, and **(b and c)** cerebellum (presented as adjusted and unadjusted for total brain volumes (TBV)). For visual purposes, we present in age (years). socioeconomic status (SES) is categorized into quartiles using ggpredict [quart2] function in R.^89^ Children from low levels of socioeconomic status had a relatively smaller global cortical surface area or cerebellum volumes and accelerated maturation of the brain compared to their developing peers. Abbreviations: SES, socioeconomic status.

**Figure 3.**
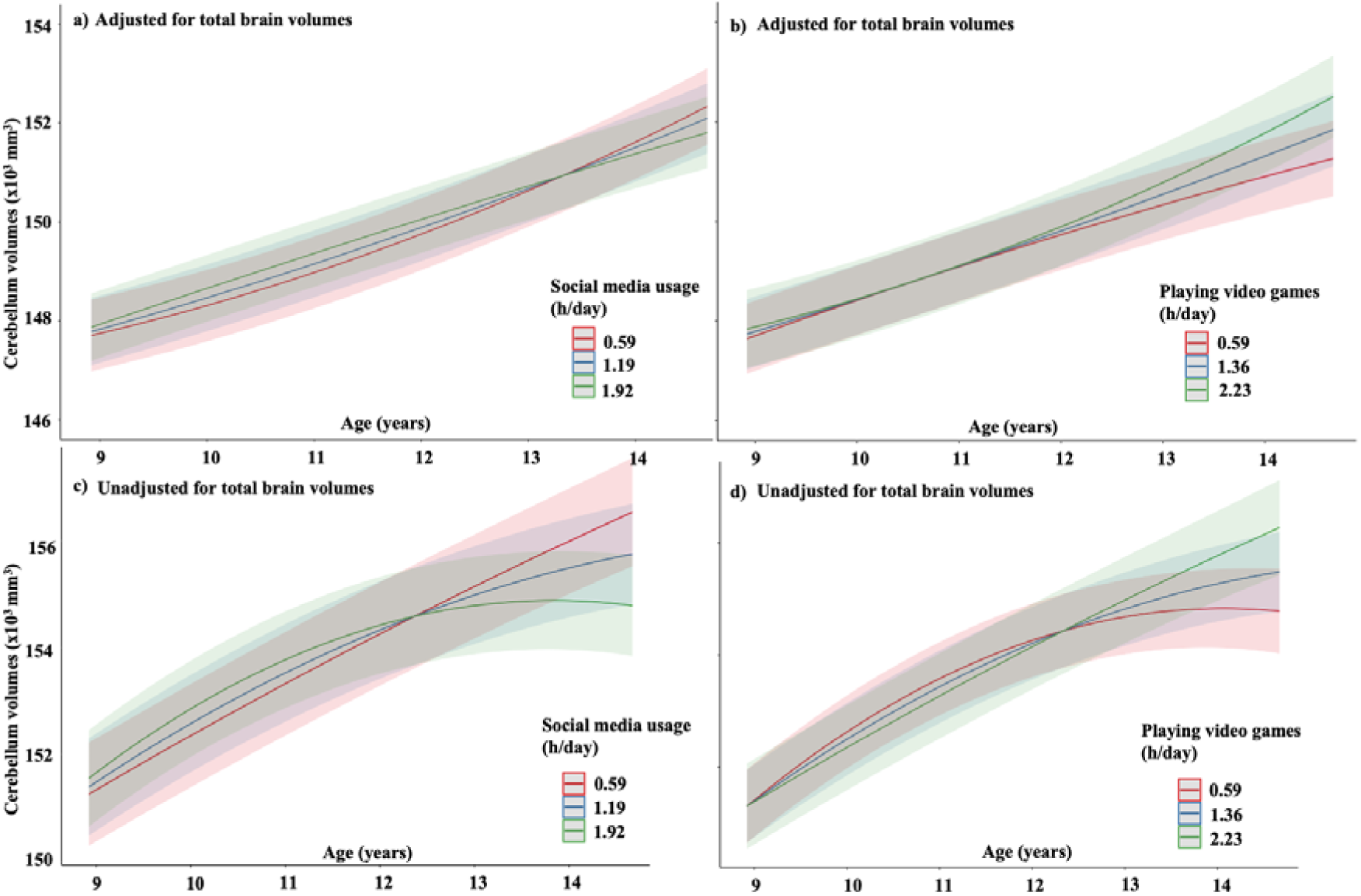
Relationship between digital media usage and cerebellum development over time. The interactions presented in **(a and c)** social media usage **(b and d)** playing video games, and time^2^ on the cerebellum development; however, they are presented in age (years) for visual purposes. Digital media usage is categorized based on quartiles using ggpredict [quart2] function in R.^89^ Children who spent a longer time on social media usage **(a and c)** had a decrease in cerebellum volume. Similar findings were seen for those who spent on mean levels. In contrast, children who spent a longer time playing video games **(b and d)** had an increase in cerebellum volume.

**Table 2.** Association between individual digital media exposure and brain outcomes in children aged 9-11 years with 4 years of follow-ups.

| Brain regions | Social media usage |  | Time (linear) |  | Time (Quadrant) |  | SES |  | cogPGS |  | Social media usage x Time |  | Social media usage x Time<sup>2</sup> |  |
| --- | --- | --- | --- | --- | --- | --- | --- | --- | --- | --- | --- | --- | --- | --- |
|  | β (SE) | p | β (SE) | P | β (SE) | p | β (SE) | P | β (SE) | P | β (SE) | P | β (SE) | P |
| Global cortical surface area (mm<sup>2</sup>) | 0.001 (0.001) | 0.67 | 0.04 (0.001) | < 0.001 | -0.03 (0.01) | <0.001 | 0.11 (0.02) | **<0.001** | 0.05 (0.02) | 0.005 | 0.001 (0.006) | 0.36 | -0.01 (0.004) | 0.004 |
| Striatum Volume (mm<sup>3</sup>) | -0.007 (0.01) | 0.49 | -0.02 (0.01) | 0.11 | -0.004 (0.005) | 0.42 | 0.01 (0.02) | 0.43 | -0.01 (0.02) | 0.40 | -0.007 (0.006) | 0.91 | 0.001 (0.003) | 0.96 |
| Cerebellum Volume (mm<sup>3</sup>) | 0.02 (0.01) | 0.10 | 0.09 (0.008) | <0.001 | 0.01 (0.005) | 0.04 | 0.05 (0.02) | 0.004 | -0.002 (0.02) | 0.98 | 0.02 (0.005) | **<0.001** | -0.02 (0.003) | **<0.001** |
|  | Playing video games |  | Time (linear) |  | Time (Quadrant) |  | SES |  | cogPGS |  | Playing video games x Time |  | Playing video games x Time<sup>2</sup> |  |
|  | β (SE) | p | β (SE) | P | β (SE) | p | β (SE) | P | β (SE) | P | β (SE) | P |  |  |
| Global cortical surface area (mm<sup>2</sup>) | -0.04 (0.01) | <0.001 | 0.03 (0.01) | 0.001 | -0.05 (0.01) | <0.001 | 0.08 (0.02) | **<0.001** | 0.03 (0.02) | 0.16 | -<0.001 (0.005) | 0.95 | 0.008 (0.003) | 0.01 |
| Striatum Volume (mm<sup>3</sup>) | -0.01 (0.01) | 0.16 | -0.001 (0.01) | 0.71 | -0.01 (0.01) | 0.10 | 0.02 (0.02) | 0.20 | -0.009 (0.02) | 0.59 | -0.01 (0.006) | 0.07 | 0.004 (0.003) | 0.16 |
| Cerebellum Volume (mm<sup>3</sup>) | 0.006 (0.009) | 0.49 | 0.12 (0.008) | <0.001 | -0.02 (0.005) | <0.001 | 0.06 (0.02) | **0.002** | -0.006 (0.02) | 0.69 | -0.008 (0.005) | 0.07 | 0.01 (0.003) | **<0.001** |
|  | Watching television |  | Time (linear) |  | Time (Quadrant) |  | SES |  | cogPGS |  | Watching television x Time |  | Watching television x Time<sup>2</sup> |  |
|  | β (SE) | p | β (SE) | p | β (SE) | p | β (SE) | P | β (SE) | P | β (SE) | P | β (SE) | P |
| Global cortical surface area (mm<sup>2</sup>) | -0.03 (0.008) | <0.001 | 0.04 (0.01) | 0.001 | -0.04 (0.006) | <0.001 | 0.08 (0.02) | **<0.001** | 0.04 (0.02) | 0.05 | -0.003 (0.004) | 0.50 | -0.001 (0.002) | 0.63 |
| Striatum | -0.02 | 0.03 | -0.03 | 0.03 | 0.003 | 0.62 | 0.02 | 0.28 | -0.005 | 0.78 | -0.003 | 0.54 | -0.002 | 0.30 |
| Volume (mm<sup>3</sup>) | (0.001) |  | (0.01) |  | (0.007) |  | (0.02) |  | (0.02) |  | (0.005) |  | (0.002) |  |
| Cerebellum | 0.007 | 0.33 | 0.08 | <0.001 | 0.006 | 0.34 | 0.03 | 0.11 | 0.03 | 0.10 | 0.01 | 0.007 | -0.006 | 0.02 |
| Volume (mm<sup>3</sup>) | (0.007) |  | (0.009) |  | (0.006) |  | (0.02) |  | (0.02) |  | (0.003) |  | (0.002) |  |

P values presented are uncorrected for multiple comparisons. The main effects that survived a number of tests were highlighted ( $p < 0.05/18 = 0.0027$ ).
Abbreviations: SES, socioeconomic status; cogPGS, polygenic scores for cognitive performance.
Model: $Y_{ij} = \beta_0 + \beta_1 cogPGS_i + \beta_2 Poly(Time, 2)_{ij} + \beta_3 Sex_i + \beta_4 Age\ at\ baseline_i + \beta_5 DM\ Usage_i + \beta_6 SES_i + \beta_7 Ancestry_i + \beta_8 (DM\ Usage_i * Poly(Time, 2)_{ij}) + \beta_9 (DM\ Usage_i * Poly(Time, 2)_{ij} * SES_i) + \beta_{10} (DM\ Usage_i * Poly(Time, 2)_{ij} * cogPGS_i) + (1 | sites) + v_{i0} + v_{i1} Poly(Time, 2)_{ij} + \varepsilon_{ij}$

No association was found between cogPGS and global CSA and the volumes of cerebellum and striatum.

There was no significant three-way interaction found between DM usage, time, and SES on any brain regions studied (i.e., global CSA, and volumes of striatum and cerebellum).

### Interaction of DM usage and time on brain development

There were multiple interactions between the average DM usage and both linear and quadratic effects of time on brain development (**Table 2)**. Only the significant interactions that survived Bonferroni corrections (p < 0.003) will be discussed below.

### Social media usage

We found a significant interaction between average social media usage and both linear and quadratic effects of time (i.e., average social media usage x time and average social media usage x time^2^, respectively) with cerebellum volume. Here, we observed a positive association of social media usage and time with cerebellum volumes for a linear term (β = 0.02) but a negative association of social media usage and time with cerebellum volumes for a quadratic term (β = −0.02) **(Table 2)**. The consequence of these effects is illustrated in **(Figure 3a and 3c)**; there is a slight difference in trajectory, which results in an earlier decline and lower volume at the last time-point.

There was no significant interaction between social media usage and time with other brain regions studied (i.e., global CSA and striatum volumes) (**Table 2)**.

### Playing video games

In contrast to social media usage on brain development, we observed a significant positive interaction between average time spent playing video games and a quadratic effect of time (but not linear) with cerebellum volume (β = 0.01) **(Table 2)**. This resulted in a trajectory with continued increase throughout the study period and a larger cerebellar volume at the last time-point **(Figure 3b and 3d)**.

There was no significant interaction between playing video games and time in other brain regions studied (i.e., global CSA and striatum volumes) (**Table 2)**.

### Watching television/videos

There was no significant interaction between watching television and time with any of the brain regions studied (i.e., global CSA, volumes of cerebellum and striatum) (**Table 2)**.

There were no significant three-way interactions between any DM usage, time, and sex with the brain structures studied. Therefore, we did not carry out separate analyses for boys and girls **(eTable 4)**.

### Additional analysis

In investigating whether DM estimates preceded changes in cerebellum volume, we observed a negative trend for high social media users with changes in cerebellum volumes (β = −0.01; p = 0.10). Conversely, there was a significant positive association for high video game users with changes in cerebellum volumes (β = 0.02; p = < 0.001) (**eTable 5)**.

We then investigated whether social media usage at T_0_ could predict total changes in cerebellum volumes (T_4_ – T_0_) and found that social media usage at T_0_ was not associated with changes in cerebellum volumes (β = −0.03; p = 0.16) (**eTable 6)**. The estimate (β = −0.03) thus represents the overall effect size of social media usage over a four-year study period.

Furthermore, after excluding time spent on video chatting or texting from social media usage, we still found that the direction of the observed effects between average social media usage on cerebellum volumes remained significant, with the same effect size (β = −0.02; p=0.002).

### Sensitivity analyses

When we excluded children who were born preterm, had low birth weight (< 2500 g), or had ADHD diagnosis, 3979 children fulfilled the criteria for eligibility, and the findings of the average social media usage or playing video games and cerebellum volumes remained significant **(eTable 7)**.

Further, our analysis was confined to children with MRI data available across all three visits (n=1462), and despite this restriction, the observed effects between average social media usage or playing video games and cerebellum volumes remained significant **(eTable 8)**.

## Discussion

In this large prospective cohort study, we studied the long-term effects of DM usage on the development of the cortex, striatum, and cerebellum across three time points spanning between mid-childhood to adolescence, in children aged 9 to 11 years. Despite our initial hypothesis, we found that DM usage did not significantly alter the development of the global CSA or striatum volume. However, children who devote more time to playing video games had a weak increase in cerebellum volume during the critical developmental window of development (β = 0.01), while those who spent more time using social media had a subtle decrease in cerebellum volume (β= −0.02). These associations persisted in subsequent analysis, even when factors such as preterm birth, lower birth weight, or those with ADHD diagnosis, were excluded, underscoring the robustness of our findings. And these associations also did not differ between the sexes. However, the effect size observed for this association was smaller than our predefined threshold of 0.05. Moreover, in analysing the accumulated differences in cerebellum volumes over four years were also very small, which is likely not of relevance to the individual. Nevertheless, this difference was accelerated during the last year **(Figure 2)**. Thus, it is relevant to conduct further research to analyse the long-term effects of social media on brain development.

The term “social media” consists of a broad spectrum of digital tools associated with social interaction, including social networking sites, text messaging applications, and video chatting. Previous studies examining the association between social media use and functional or neural outcomes in both children and adolescents have often either combined all these digital tools under the umbrella term “social media use”,^4,28^ or scrutinized them separately, distinguishing between social media platforms (e.g., Facebook) and social communication tools (e.g., text messaging) in their analyses.^29,30^ Consistent with previous studies we first investigated the effect of social media usage on brain development by combining all these digital tools. We specifically included activities related to social media platforms and studied their singular long-term effect on cerebellum development. Even in this refined analysis, we still observed a persistent weak negative effect of social media usage on cerebellum volumes.

If the negative developmental trend for the cerebellum persists, it might be of significant concern, particularly considering that adolescence serves as the period when many psychiatric disorders have their onset.^31,32^ Moreover, consistent findings report an association between cerebellum abnormalities with various psychiatric disorders, such as depression and anxiety disorders.^33^ In addition, the cerebellum is a core component of the neural circuitry underpinning many cognitive deficits associated with ADHD, including working memory, response inhibition, attention shifting, and processing of rewards and temporal information.^34–37^

The cerebellum is sensitive to environmental exposures both prenatally, as demonstrated by studies of maternal alcohol, maternal diabetes, hypoxia, and postnatal glucocorticoid exposure,^38,39^ and postnatally.^40^ In our study, we observed that children from lower SES quartiles had smaller cerebellum volumes, providing further for the susceptibility of the cerebellum structure to environmental factors.^41,42^ The transition from childhood to adolescence represents a critical developmental phase characterized by hormonal and physiological changes, including myelination, strengthening of synapses, and selective pruning of neurons and connections. Social media users often contend with constant distractions, which can significantly impact their behavior, leading to inattention symptoms.^43^ Additionally, these users can become easily diverted from tasks like reading or homework, etc. Moreover, the use of social media necessitates continual response to stimuli, decision-making, and the execution of motor movements, among various other cognitive and behavioral tasks. Previous studies on social media usage have consistently reported negative effects on life satisfaction,^7^ overall well-being,^44^ and depressive symptoms,^45^ among adolescents. Based on these observations, one might speculate that a distinct window of susceptibility to emotion and frequent shifts in task stimuli might be key contributing factors to the observed decrease in cerebellum volumes. At the neuronal level, this could reflect the acceleration of the natural process of synaptic pruning and changes in myelination among high social media users, which would then appear as a decrease in cerebellum volume at a later time point.

Consistent with prior research,^46,47^ we observed an inverted U-shaped trajectory in the development of the cortex during mid-childhood and adolescence, with girls reaching their peak earlier than boys. These findings align with histological studies suggesting continued myelination and reduction in synaptic density during adolescence.^48,49^ At a microscopic level, cortical maturation involves synaptic overproduction in childhood, followed by selective elimination and strengthening of connections later in development.^50^ During these stages of development, environmental exposure might guide selective synapse elimination in adolescence.^51,52^ Supporting this notion, we found that children from lower SES quartiles exhibited smaller global CSA across development compared to their peers.

Although this is a longitudinal study with a large number of participants, the study has some notable limitations. First, this is an observational study, and therefore, we cannot establish causal inference. However, we adjusted for most of the covariates such as age, sex, SES, and genetics. Additionally, to mitigate selection bias, we ensured the inclusion of only one child per family. Second, the estimated time spent on various DM types was self-reported, introducing potential recall or accuracy bias. Nevertheless, it should be noted that studies have reported high test and retest reliability of self-reported behaviors among adolescents.^53^ Third, the survey questionnaire utilized to capture DM usage from T_2_ visits onwards was modified compared with T_0_ or T_1_ visits in response to technological advancements and the heightened usage of DM among adolescents. However, we harmonized the survey questionnaires from the T_2_ visit onwards to maintain consistency with the earlier visits. Fourth, the response measure for the survey questionnaire in both T_0_ and T_1_ visits was set between ‘0 and 4+ hours’; this is one of the major drawbacks of the ABCD questionnaire. For example, a child who spent four hours engaged in video games or using social media would receive the same score as a child who spent 12 hours, despite the significant difference in their exposure. Fifth, the ABCD questionnaire failed to capture information regarding the timing of DM usage, either during the day or night, thus impeding the exploration of the potential effects of bedtime DM usage on brain development. Finally, the survey questionnaires used in this study failed to capture any information regarding the genre of video games. Given that different activities and actions of video gaming may exert distinct impacts on brain development.

In summary, DM usage, particularly playing video games, does not alter cortical brain development during the four-year window, but social media usage is weakly associated with a decrease in cerebellum volumes, a trend that was accelerated at later time-points. These findings should be continued by longer follow-up, and more detailed documentation of DM usage, but is a cause for concern regarding the usage of social media in children and adolescents.

## Methods

### Participants

The neuroimaging and behavioral data used in this study were obtained from the ABCD Study (data release 5.0; https://abcdstudy.org/; http://doi.org/10.15154/1523041), a longitudinal cohort of 11,875 children born between 2005 and 2009. These children were enrolled at ages 9-11 years from 21 research sites across the U.S. between 2016 and 2018,^54^ with the intention of following them for a period of at least 10 years. This recruitment cohort closely matches the sociodemographic composition of the US population of 9-11-year-old children. Most of the children were enrolled through local elementary and charter schools at each data-collection site. A smaller portion was recruited through community outreach and word-of-mouth referrals outside of the school setting. Twins were identified and recruited from birth registries.^55,56^

During each visit, children accompanied by a parent/guardian, completed a series of measures. These included neurocognitive tests, mental and physical health questionnaires, environmental exposure data collection, providing biological specimens, and participating in brain imaging.^54,57–61^ All were asked for an in-person assessment session for self- or parent-report of mentioned behavioral measures and for biological specimen collections once a year, with brain imaging conducted biannually. For this study, we used data collected between September 2016 and January 2022, which included baseline (T_0_), one-year follow-up (T_1_), two-year follow-up (T_2_), three-year follow-up (T_3_), and four years follow-up (T_4_).^60,61^ Children were excluded if they were born extremely preterm (< 28 weeks of gestation) or had birth weight (< 1200 g), were not proficient in English, had any neurological problems, had a history of seizures, or had a contraindication to undergo brain MRI scans. All children and their parents/guardians provided informed written consent/assent for participation, and the central Institutional Review Board at the University of California, San Diego approved the study protocols. All the research methods were performed in accordance with the relevant guidelines and regulations.

Children who did not have relevant data on either SES, genetics, DM usage; or neuroimaging were excluded from the present study. Additionally, the ABCD cohort included twins and siblings, therefore we randomly selected one child per family to eliminate this source of bias.

### Neuroimaging

Children underwent brain MRI scans on 3-Tesla scanner platforms (Siemens Prisma, Philips, or General Electric 750) using a standard adult-sized head coil at three different time points over a span of four years (i.e., T_0_, two years later (T_2_), and four years later (T_4_)). A standardized protocol for scanning was used to harmonize the scanning sites and MRI scanners. Three-dimensional T1-weighted images (1-mm isotropic) were acquired using a magnetization-prepared rapid acquisition gradient-echo (MP-RAGE) sequence and processed using FreeSurfer software (version 5.3.0).^62^ All the pre-processed images were quality-checked according to the ABCD protocol, as described earlier,^39^ and children with excessive head motion or poor image quality were excluded from the current study. In brief, all the imaging data and FreeSurfer outputs were evaluated by the ABCD Data Analysis, Informatics, and Resource Center (DAIRC) image processing pipeline for real-time motion detection and correction.^54,57^ In addition, FreeSurfer output was rated manually by a trained technician for the following errors: motion, homogeneity, white-matter underestimation, pial overestimation, and magnetic susceptibility artifacts; and were rated from 0 to 3 (0=absent, 1=mild, 2=moderate, and 3=severe). As per the ABCD study recommendation, we excluded children with poor scan quality, did not pass manual quality check, or with any incidental findings.

The Destrieux atlas was used to calculate total brain volumes and global cortical surface area (CSA), while the ASEG atlas was used to segment both striatum and cerebellum volumes.^57,62,63^

### Covariates

#### Socioeconomic status

SES was defined as the first principal component from a probabilistic principal component analysis (PCA), capturing 65% of the variance in total household income, highest parental education, and neighbourhood quality. Children missing more than one of these SES measures were excluded. Household income was determined by the combined annual income of all family members over the past 12 months, categorized as less than $49,999 (1), $50,000–74,999 (2), $75,000–99,999 (3); $100,000–199,999 (4); and greater than $200,000 (5). Parental education was categorized into middle school or less (1), some high school (2), high school graduate (3), some college/associate degree (4), bachelor’s degree (5), master’s degree (6), or professional degree (7). The neighbourhood quality was determined using the area deprivation index, calculated from the American Community Survey using the address of the primary residency.^64^ The SES composite and each subcomponent were normalized (mean=0, SD=1).

### Polygenic Score Derivation and Analyses

#### Genotyping, quality control, and imputation

Saliva samples were collected from all the children during the T_0_ visit and genotyped using Rutgers University Cell and DNA repository using the Smokescreen array consisting of 646,247 genetic variants.^65^

Quality control, imputation, and genetic PCA were performed by the National Bioinformatics Infrastructure Sweden (NBIS). The following pre-processing steps were conducted. Briefly, single nucleotide polymorphisms (SNPs) with call rates < 98% or minor allele frequencies (MAFs) < 1% were excluded before imputation. Individuals with high rates of missingness (> 2%) and absolute autosomal heterozygosity > 0.2 were excluded, resulting in 10,069 children and 430,622 genetic variants. Haplotypes were prephased using SHAPEIT2, and genetic markers were imputed using IMPUTE4 software.

We utilized the 1000 Genomes haplotypes—Phase 3 integrated variant set release in NCBI build 37 (hg19) coordinates as reference populations. This dataset consists of 2504 samples and 5008 haplotypes from Europeans, Africans, East Asians, Southern Asians, and Americans (https://mathgen.stats.ox.ac.uk/impute/1000GP_Phase3.html). We used this imputation since it provides better concordance in diverse human populations.^66,67^ After that, genotypes with an INFO score < 0.3 or MAF < 0.001% were excluded, which yielded 40,637,119 SNPs in a total of 10,069 children.

The PCA module, as implemented in RICOPILI,^68^ was used to check for outliers and control population structure. SNPs were pruned so that there was little linkage disequilibrium (LD) between SNPs (R2 < 0.2, 200 SNP window: Plink–indep-pairwise 200 100 0.2). LD pruning was repeated until 100 K SNPs were reached. The resulting SNPs were then projected into the PCA.^69,70^ We utilized the first 20 principal components (20PCs) from the genetic PCA.

#### cogPGS calculation

We created polygenic scores for cognitive performance (cogPGS) in each child using PRSice-2,^71^ which involved summing the effect sizes of thousands of SNPs (weighted by the presence of effect alleles in each child). These SNPs were discovered by large genome-wide association studies (GWAS) on educational attainment, mathematical ability, and general cognitive ability.^72^ Details regarding the effect sizes and p values of their SNPs can be assessed through the Social Science Genetics Association Consortium (https://www.thessgac.org/data).

We utilized the data provided by the consortium from a multitrait analysis of GWAS,^73^ which, in our case, represents a joint polygenic score focused on a GWAS of cognitive performance and complemented by information from a GWAS on educational attainment, a GWAS on the highest-level math class completed, and a GWAS on self-reported math ability. This joint analysis is ideal because pairwise genetic correlations of these traits were high,^72^ and these GWAS had hundreds of thousands of individuals. Such a large sample size allows new studies to detect effects in samples of a few hundred individuals with 80% statistical power.

To construct the cogPGS, we performed clumping and pruning to remove nearby SNPs that are correlated with each other. The clumping sliding window was 250 kb, with the linkage disequilibrium clumping set to r^2^ > 0.25. We included the weightings of all SNPs, regardless of their p-value from the GWAS (p = 1.00 threshold), resulting in 5255 SNPs. Finally, we normalized (mean=0, SD=1) the cogPGS to fairly compare their effects on different phenotypes. For the present study, we used cogPGS to reflect the genetic predisposition of cognitive performance and included 20 genetic principal components (PCs) to account for the possibility of population stratification within the Add Health European-ancestry subsample in the same model.

### Exposure

#### Digital media usage

The estimated time spent on individual DM usage (i.e., using social media, playing video games, or watching television/videos) was assessed at all annual visits (i.e., T_0_, one year later (T_1_), T_2_, three years later (T_3_), and T_4_) using the self-reported Youth Screen Time Survey.

#### Self-report survey

At each visit, children reported the number of hours they spent on a typical weekday (i.e., Monday to Friday during the school year and holiday/school breaks) as well as weekend days (i.e., Saturday and Sunday). These hours were categorized by device, media platform, or activity excluding the number of hours spent on school-related work. Specifically, they reported the number of hours dedicated to the following activities:

1. watching television or movies,
2. watching videos (e.g., YouTube),
3. playing video games on a computer, console, phone, or another device (e.g., Xbox, PlayStation, iPad),
4. Texting on a cell phone, tablet, or computer (e.g., Google Chat, WhatsApp),
5. Visiting social networking sites (e.g., Facebook, Twitter, Instagram), and
6. Using video chat (e.g., Skype, FaceTime).

To be consistent with our earlier study,^4^ we categorized DM usage as follows: (a) using social media (4+5+6), (b) playing video games (3), or (c) watching television/videos (1+2). The response options included were none - ‘0’, < 30 minutes - ‘0.25’, 30 minutes – ‘0.5’, 1 hour – ‘1’, 2 hours – ‘2’, 3 hours – ‘3’, or > 4 hours – ‘4’.

To calculate the average hours spent per day for individual DM usage, the following formula was used: [(total number of hours spent on a weekday * 5) + (the total number of hours on a weekend day * 2)]/7.

For both the T_0_ and T_1_ visits, data were collected using the same categorical scale as described above. However, starting from T_2_, modifications were made to the Youth Screen Time Survey to accommodate the increasing DM usage among school-aged children. The time spent watching television was changed into ‘watching or streaming videos or movies’, while watching videos (such as YouTube) was changed into ‘watching or streaming videos or live streaming (such as YouTube, Twitch)’. Then, these categories were merged into a single category named ‘watching television/videos’. ‘Video chatting, visiting social media apps, and texting cell phone’ were combined into a broader category called ‘using social media’. The activities ‘editing photos and videos’ and ‘searching or browsing the internet’ were excluded as they were not present in the T_0_ data. Playing video games was further divided into two subcategories, i.e., ‘time spent on single-player’ and ‘time spent on multi-player’, which were combined as ‘playing video games’.

Additionally, the response format was changed from categorical to continuous, with response options including 0 minutes, 15 minutes, 30 minutes, 45 minutes, 1 hour, 1.5 hours, 2 hours, 2.5 hours, 3 hours, and every additional hour up until 24 hours.

To ensure consistency across all time points, we standardized the T_2_, T_3_, and T_4_ visit data to align with the T_0_ and T_1_ visit data. As a result, the data from the T_2_, T_3_, and T_4_ visits were recoded to match the categories used in the T_0_ and T_1_ visits. The recoding involved transforming the continuous response options into the following categories: none -‘0’, < 30 minutes -‘0.25’, 30 minutes – ‘0.5’, 1 hour – ‘1’, 1.15 hours – ‘1.25’, 1.30 hours – ‘1.5’, 2 hours – ‘2’, 2.15 hours – ‘2.25’, 2.30 hours – ‘2.5’, 3 hours – ‘3’, 3.15 hours – ‘3.25’, 3.30 hours – ‘3.5’, and > 4 hours – ‘4’.

There were good test-retest correlations between individual DM usage across different time points, with r-values ranging from 0.24 to 0.56 **(eFigure 1)**.

#### Parent-reported survey

Caregivers/parents were asked to report the number of hours spent by their child on a typical weekday and weekend day engaging in total on watching television, shows or videos, texting or chatting, playing games, or visiting social networking sites (Facebook, Twitter, Instagram), excluding the number of hours spent on school-related work during T_0_ and T_1_ visits. Parents provided the total estimated time spent on these activities in both hours and minutes for weekdays and weekends. To calculate the average hours of screen time per day, we used the following: [(total number of hours spent on a weekday * 5) + (the total number of hours on a weekend day * 2)]/7.

Furthermore, we assessed the agreement between caregivers/parents and child reports regarding the estimated amount of time spent on total screen activities (i.e., watching television/videos, playing video games, or engaging in social media) during the T_0_ visit, using a correlation coefficient and found it to be 0.37, indicating fair agreement between caregivers/parents and children. To obtain the child’s report on the estimated screen time at T_0_, we summed the time spent watching television/videos, playing video games, or using social media.

We opted to use the self-reported surveys completed by the children rather than relying on caregivers/parents.^74,75^ Since caregivers/parents may not be fully aware of specific types of DM used by their children, including those aged 9-11 years and older, who often use DM without supervision, such as in their bedrooms at night. Consequently, children may provide more accurate reports of their estimated time spent on each type of DM usage. There is also substantial evidence showing that children as young as six years old can reliably report on their own health.^75^

In light of the COVID-19 lockdown, it is probable that these children could spend more time using DM than anticipated at T_0_. This effect was more pronounced in a US-based study, which reported a two-fold increase in the estimated time spent on DM usage during the COVID-19 lockdown compared to the pre-pandemic period.^76^ Therefore, to account for an increase in estimated time spent using DM among children between T_0_ and T_4_, we used the average estimated time spent for individual DM usage, rather than relying solely on data from either T_0_ or T_4_ for the longitudinal analyses. The average estimated time spent for individual DM usage was calculated by averaging the estimated time spent for each type of DM usage across all time points.

### Outcomes

These predefined outcomes included the global CSA and the volumes of the striatum and cerebellum. We defined the striatum by combining the volumes of the caudate nucleus, putamen, and accumbens. As for the cerebellum, we combined the volumes of both grey and white matter structures of the cerebellum. Both striatum and cerebellum volumes were adjusted for the total brain volumes. In these analyses, we considered both the left and right hemispheres together.

### Statistical analysis

Descriptive statistics including means and standard deviations (SDs) were calculated.

The first research question aimed to assess whether individual DM usage altered (i.e., increased or decreased) brain development over four years.

To address this question, we first inspected the developmental trends of brain structures (i.e., global CSA, cerebellum, and striatum) between mid-childhood and early adolescence, which are not always linear. Earlier studies on brain development have reported both linear and quadratic trends between childhood and adolescence.^77–80^ To do so, we compared the default linear model to a complex quadratic model to identify whether adding the quadratic age effect significantly improved the goodness of fit for the global CSA, cerebellum, and striatum. In both these models, we adjusted for SES, polygenic scores cogPGS, and 20PCs. We assessed the fit of the models based on the Akaike Information Criterion (AIC) and Bayesian Information Criterion (BIC). The model with lower AIC and BIC values was considered a better fit (at least by 10 points less than the other model) **(eTable 1)**.^81^ The log-likelihood ratio test (χ^2^) was additionally run to confirm the results.

When we examined the models, the quadratic model fitted the data well and was subsequently used for further analysis. In addition, age-related change in the peak location along with sex effect was assessed. Peak age for each brain structure was calculated using the first derivative of the quadratic equations.

We constructed a quadratic mixed-effect model to investigate the relationship between individual DM usage and brain structures over time. The model (equation 1) was adjusted for various factors: age at baseline (mean-centered to reduce multicollinearity), SES, cogPGS, 20 PCs, and sex assigned at birth as fixed effects, and study sites were included as random effects.

To test the long-term effect of DM usage on brain development with time (as outcomes of interest), we included a two-way interaction with average DM usage and time as both linear and quadratic terms (i.e., average DM usage x Time; average DM usage x Time^2^). Furthermore, to account for SES and cogPGS effects on brain development over time, we included three-way interactions in the same model (i.e., for SES, average DM usage x Time x SES; and average DM usage x Time^2^ x SES; for cogPGS, average DM usage x Time x cogPGS; and average DM usage x Time^2^ x cogPGS). Both the intercepts and the slopes were used as random-effects terms, allowing children to start at different levels of surface area/volumes. The ‘lmer’ function of package lme4 in R software was used to fit the model, and the restricted maximum likelihood method was used to estimate the model parameters.^82,83^

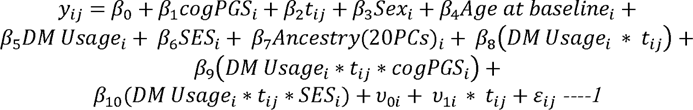

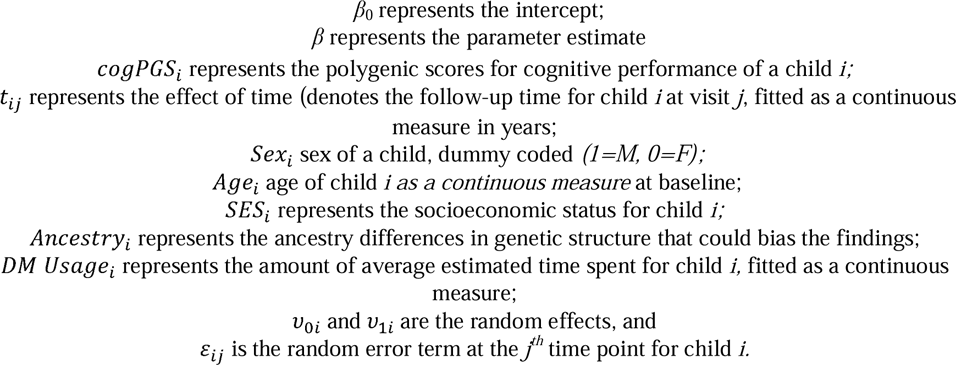

To determine the effect of sex-related differences on the relationships between DM usage and brain development, we added an interaction effect of sex (i.e., average DM usage x Time x sex; and average DM usage x Time^2^ x sex) to the pre-existing model (equation 2).

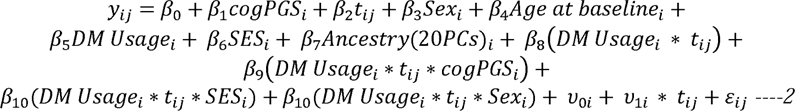

Considering the numerous statistical tests conducted, the Bonferroni corrections were applied to control for Type-1 error.^84^ In total, we performed three individual DM usage models (i.e., using social media, playing video games, watching television/videos) for three brain structures analyzed in the overall cohort as well as for sex, resulting in a total of 18 tests. *P* < 0.003 was considered statistically significant.

An additional analysis was conducted to investigate whether the estimates of DM preceded changes in cerebellum volume. A linear model was employed to ascertain whether the average social media usage of the first two time points (i.e., (T_0_ + T_1_)/2) could predict later changes in cerebellum volumes between T_2_ and T_4_, while adjusting for the aforementioned covariates (i.e., age at baseline, SES, cogPGS, 20 PCs, and sex assigned at birth as fixed effects, and study sites as random effects). Subsequently, the same analysis was repeated using the average time spent playing video games during the first two time points in cerebellum volumes. We then investigated whether social media usage at T_0_ could predict the changes in cerebellum volumes (T_4_ – T_0_) over the study period, while adjusting for prespecified covariates as mentioned above. In addition, we explored whether excluding time spent on video chatting or texting from social media usage would alter the results **(equation 1**; **Table 2).**

We ran multiple robustness tests to validate our findings and they were uncorrected. Firstly, we excluded children who were born preterm (< 37 weeks), had low birth weight (< 2500 g) or had a diagnosis of ADHD. Those born preterm or with low birth weight tend to have altered developmental trajectories.^85–87^ Similarly, children with ADHD have delayed maturation, which might affect our findings.^88^ Secondly, we restricted our analysis by including children with MRI data for all three-time points.

The gestation length and birth weight of each child were reported by caregivers/parents through a self-reported questionnaire. The presence of ADHD symptoms in the child, whether in the past or currently, was assessed through caregivers/parents reports using the computerized Kiddie-Structured Assessment for Affective Disorders and Schizophrenia (KSADS) during the T_0_ visit. This tool is based on a well-studied and validated tool, both in research and clinical settings. Diagnoses of ADHD were made in accordance with *DSM-5* criteria, which require an endorsement of six or more symptoms of inattention or hyperactivity-impulsivity.

## Authors contribution

Samson Nivins: Conceptualization, Writing – original draft, Investigation, Methodology, Formal analysis, Visualization. Bruno Sauce, Nicholas Judd: Conceptualization, Methodology, Writing – review & editing. Magnus Liebherr: Conceptualization, Writing – review & editing. Torkel Klingberg: Conceptualization, Methodology, Writing – critical review, insights, & editing, Funding acquisition.

## Data Availability statement

The data used for the analyses presented in this paper are from the Adolescent Brain Cognitive Development (ABCD) Study [https://abcdstudy.org; NIMH Data Archive (NDA)]. Data can be accessed by directly applying to the NDA.

## Code Availability

The code to replicate all analysis described in this manuscript can be found here:

https://github.com/samniv/ScreenTime-and-Brain/tree/main

## Acknowledgments

Data used in the preparation of this article were obtained from the Adolescent Brain Cognitive Development (ABCD) Study (https://abcdstudy.org), held in the NIMH Data Archive (NDA). The ABCD study was supported by the National Institutes of Health and additional federal partners under award numbers U01DA041022, U01DA041025, U01DA041028, U01DA041048, U01DA041089, U01DA041093, U01DA041106, U01DA041117, U01DA041120, U01DA041134, U01DA041148, U01DA041156, U01DA041174, U24DA041123, and U24DA041147. A full list of supporters is available at https://abcdstudy.org/?s=nIH+collaborators. A listing of participating sites and a complete listing of the study investigators can be found at https://abcdstudy.org/principal-investigators.html. The ABCD consortium investigators designed and implemented the study and/or provided data but did not necessarily participate in the analysis or writing of this report. This manuscript reflects the views of the authors and may not reflect the opinions or views of the NIH or ABCD consortium investigators.

## Additional Information

The ABCD data repository grows and changes over time. The ABCD data used in this report came from ABCD (data release 5.0; https://abcdstudy.org/; http://doi.org/10.15154/1523041).

## References

1. Media CS. The Common Sense Census: Media Use by Tweens and Teens, 2019. Accessed June, 2022.

2. Lissak G. Adverse physiological and psychological effects of screen time on children and adolescents: Literature review and case study. Environmental research. 2018;164:149–157. doi:10.1016/j.envres.2018.01.015

3. Kostyrka-Allchorne K, Cooper NR, Simpson A. The relationship between television exposure and children’s cognition and behaviour: A systematic review. Developmental review. 2017;44:19–58. doi:10.1016/j.dr.2016.12.002

4. Sauce B, Liebherr M, Judd N, Klingberg T. The impact of digital media on children’s intelligence while controlling for genetic differences in cognition and socioeconomic background. Scientific reports. 2022;12(1):7720–7720. doi:10.1038/s41598-022-11341-2

5. Thorell LB, Burén J, Ström Wiman J, Sandberg D, Nutley SB. Longitudinal associations between digital media use and ADHD symptoms in children and adolescents: a systematic literature review. European Child & Adolescent Psychiatry. 2022:1–24.

6. Wu D, Dong X, Liu D, Li H. How Early Digital Experience Shapes Young Brains During 0-12 Years: A Scoping Review. Early Education and Development. 1–37. doi:10.1080/10409289.2023.2278117

7. Orben A, Przybylski AK, Blakemore SJ, Kievit RA. Windows of developmental sensitivity to social media. Nat Commun. Mar 28 2022;13(1):1649. doi:10.1038/s41467-022-29296-3

8. Soares PSM, de Oliveira PD, Wehrmeister FC, Menezes AMB, Gonçalves H. Screen time and working memory in adolescents: A longitudinal study. Journal of Psychiatric Research. 2021/05/01/ 2021;137:266–272. 10.1016/j.jpsychires.2021.02.066

9. Takeuchi H, Taki Y, Hashizume H, et al. The Impact of Television Viewing on Brain Structures: Cross-Sectional and Longitudinal Analyses. Cerebral Cortex. 2015;25(5):1188–1197. doi:10.1093/cercor/bht315

10. Takeuchi H, Taki Y, Hashizume H, et al. Impact of videogame play on the brain’s microstructural properties: cross-sectional and longitudinal analyses. Molecular Psychiatry. 2016/12/01 2016;21(12):1781-1789. doi:10.1038/mp.2015.193

11. Takeuchi H, Taki Y, Asano K, et al. Impact of frequency of internet use on development of brain structures and verbal intelligence: Longitudinal analyses. Hum Brain Mapp. Nov 2018;39(11):4471–4479. doi:10.1002/hbm.24286

12. Miller J, Mills KL, Vuorre M, Orben A, Przybylski AK. Impact of Digital Screen Media Activity on Functional Brain Organization in Late Childhood: Evidence from the ABCD Study. Cortex. 2023/10/19/ 2023;10.1016/j.cortex.2023.09.009

13. Dick AS, Lopez DA, Watts AL, et al. Meaningful associations in the adolescent brain cognitive development study. Neuroimage. Oct 1 2021;239:118262. doi:10.1016/j.neuroimage.2021.118262

14. Funder DC, Ozer DJ. Evaluating effect size in psychological research: Sense and nonsense. Advances in Methods and Practices in Psychological Science. 2019;2(2):156–168.

15. Abelson RP. A variance explanation paradox: When a little is a lot. Psychological Bulletin. 1985;97(1):129–133. doi:10.1037/0033-2909.97.1.129

16. Anvari F, Kievit R, Lakens D, et al. Not All Effects Are Indispensable: Psychological Science Requires Verifiable Lines of Reasoning for Whether an Effect Matters. Perspect Psychol Sci. Mar 2023;18(2):503–507. doi:10.1177/17456916221091565

17. Schnack HG, Van Haren NEM, Brouwer RM, et al. Changes in Thickness and Surface Area of the Human Cortex and Their Relationship with Intelligence. Cerebral cortex (New York, NY 1991). 2015;25(6):1608–1617. doi:10.1093/cercor/bht357

18. Skranes J, Løhaugen GCC, Martinussen M, Håberg A, Brubakk A-M, Dale AM. Cortical surface area and IQ in very-low-birth-weight (VLBW) young adults. Cortex. 2013;49(8):2264–2271. doi:10.1016/j.cortex.2013.06.001

19. Walhovd KB, Krogsrud SK, Amlien IK, et al. Neurodevelopmental origins of lifespan changes in brain and cognition. Proceedings of the National Academy of Sciences. 2016;113(33):9357–9362. doi:doi:10.1073/pnas.1524259113

20. Fjell AM, Westlye LT, Amlien I, et al. High-Expanding Cortical Regions in Human Development and Evolution Are Related to Higher Intellectual Abilities. Cerebral Cortex. 2013;25(1):26–34. doi:10.1093/cercor/bht201

21. Williams CM, Peyre H, Ramus F. Brain volumes, thicknesses, and surface areas as mediators of genetic factors and childhood adversity on intelligence. Cerebral Cortex. 2022;33(10):5885–5895. doi:10.1093/cercor/bhac468

22. Vuoksimaa E, Panizzon MS, Chen CH, et al. The Genetic Association Between Neocortical Volume and General Cognitive Ability Is Driven by Global Surface Area Rather Than Thickness. Cereb Cortex. Aug 2015;25(8):2127–37. doi:10.1093/cercor/bhu018

23. Judd N, Sauce B, Wiedenhoeft J, et al. Cognitive and brain development is independently influenced by socioeconomic status and polygenic scores for educational attainment. Proceedings of the National Academy of Sciences - PNAS. 2020;117(22):12411–12418. doi:10.1073/pnas.2001228117

24. Kühn S, Gallinat J, Mascherek A. Effects of computer gaming on cognition, brain structure, and function: a critical reflection on existing literature. Dialogues in clinical neuroscience. 2019;21(3):319–330. doi:10.31887/DCNS.2019.21.3/skuehn

25. Pujol J, Fenoll R, Forns J, et al. Video gaming in school children: How much is enough? Annals of neurology. 2016;80(3):424–433. doi:10.1002/ana.24745

26. Bethlehem RAI, Seidlitz J, White SR, et al. Brain charts for the human lifespan. Nature. 04 2022;604(7906):525–533. doi:10.1038/s41586-022-04554-y

27. Bagot KS, Tomko RL, Marshall AT, et al. Youth screen use in the ABCD® study. Dev Cogn Neurosci. Sep 1 2022;57:101150. doi:10.1016/j.dcn.2022.101150

28. Hamilton JL, Hutchinson E, Evankovich MR, Ladouceur CD, Silk JS. Daily and average associations of physical activity, social media use, and sleep among adolescent girls during the COVID-19 pandemic. J Sleep Res. Feb 2023;32(1):e13611. doi:10.1111/jsr.13611

29. Miller J, Mills KL, Vuorre M, Orben A, Przybylski AK. Impact of digital screen media activity on functional brain organization in late childhood: Evidence from the ABCD study. Cortex. 2023/12/01/ 2023;169:290–308. 10.1016/j.cortex.2023.09.009

30. Mougharbel F, Chaput JP, Sampasa-Kanyinga H, et al. Heavy social media use and psychological distress among adolescents: the moderating role of sex, age, and parental support. Front Public Health. 2023;11:1190390. doi:10.3389/fpubh.2023.1190390

31. Solmi M, Radua J, Olivola M, et al. Age at onset of mental disorders worldwide: large-scale meta-analysis of 192 epidemiological studies. Molecular Psychiatry. 2022/01/01 2022;27(1):281–295. doi:10.1038/s41380-021-01161-7

32. McGrath JJ, Al-Hamzawi A, Alonso J, et al. Age of onset and cumulative risk of mental disorders: a cross-national analysis of population surveys from 29 countries. The Lancet Psychiatry. 2023/09/01/ 2023;10(9):668–681. 10.1016/S2215-0366(23)00193-1

33. Phillips JR, Hewedi DH, Eissa AM, Moustafa AA. The cerebellum and psychiatric disorders. Front Public Health. 2015;3:66. doi:10.3389/fpubh.2015.00066

34. Durston S, van Belle J, de Zeeuw P. Differentiating frontostriatal and fronto-cerebellar circuits in attention-deficit/hyperactivity disorder. Biol Psychiatry. Jun 15 2011;69(12):1178–84. doi:10.1016/j.biopsych.2010.07.037

35. Noreika V, Falter CM, Rubia K. Timing deficits in attention-deficit/hyperactivity disorder (ADHD): evidence from neurocognitive and neuroimaging studies. Neuropsychologia. Jan 2013;51(2):235–66. doi:10.1016/j.neuropsychologia.2012.09.036

36. Strick PL, Dum RP, Fiez JA. Cerebellum and nonmotor function. Annu Rev Neurosci. 2009;32:413–34. doi:10.1146/annurev.neuro.31.060407.125606

37. Toplak ME, Dockstader C, Tannock R. Temporal information processing in ADHD: findings to date and new methods. J Neurosci Methods. Feb 15 2006;151(1):15–29. doi:10.1016/j.jneumeth.2005.09.018

38. Limperopoulos C, Soul JS, Gauvreau K, et al. Late gestation cerebellar growth is rapid and impeded by premature birth. Pediatrics. Mar 2005;115(3):688–95. doi:10.1542/peds.2004-1169

39. Nivins S, Klingberg T. Effects of prenatal exposure to maternal diabetes mellitus on deep grey matter structures and attention deficit hyperactivity disorder symptoms in children. Acta Paediatrica. 2023/07/01 2023;112(7):1511–1523. 10.1111/apa.16756

40. Cavanagh J, Krishnadas R, Batty GD, et al. Socioeconomic Status and the Cerebellar Grey Matter Volume. Data from a Well-Characterised Population Sample. The Cerebellum. 2013/12/01 2013;12(6):882–891. doi:10.1007/s12311-013-0497-4

41. Giedd JN, Schmitt JE, Neale MC. Structural brain magnetic resonance imaging of pediatric twins. Human brain mapping. 2007;28(6):474–481.

42. Gilmore JH, Schmitt JE, Knickmeyer RC, et al. Genetic and environmental contributions to neonatal brain structure: a twin study. Human brain mapping. 2010;31(8):1174–1182.

43. Samson Nivins MAM, Torkel Klingberg. Screen time use and longitudinal effects on ADHD symptoms in children. PsyArXiv2024.

44. Booker CL, Kelly YJ, Sacker A. Gender differences in the associations between age trends of social media interaction and well-being among 10-15 year olds in the UK. BMC Public Health. Mar 20 2018;18(1):321. doi:10.1186/s12889-018-5220-4

45. Kelly Y, Zilanawala A, Booker C, Sacker A. Social Media Use and Adolescent Mental Health: Findings From the UK Millennium Cohort Study. EClinicalMedicine. Dec 2018;6:59–68. doi:10.1016/j.eclinm.2018.12.005

46. Giedd JN, Blumenthal J, Jeffries NO, et al. Brain development during childhood and adolescence: a longitudinal MRI study. Nature neuroscience. 1999;2(10):861–863.

47. Lenroot RK, Gogtay N, Greenstein DK, et al. Sexual dimorphism of brain developmental trajectories during childhood and adolescence. Neuroimage. Jul 15 2007;36(4):1065–73. doi:10.1016/j.neuroimage.2007.03.053

48. Huttenlocher PR. Synaptic density in human frontal cortex-developmental changes and effects of aging. Brain Res. 1979;163(2):195–205.

49. Yakovlev PI, Lecours A-R, Minkowski A, _Davis F. Regional development of the brain in early life. 1967.

50. Neniskyte U, Gross CT. Errant gardeners: glial-cell-dependent synaptic pruning and neurodevelopmental disorders. Nat Rev Neurosci. Nov 2017;18(11):658–670. doi:10.1038/nrn.2017.110

51. Kleim JA, Lussnig E, Schwarz ER, Comery TA, Greenough WT. Synaptogenesis and Fos expression in the motor cortex of the adult rat after motor skill learning. Journal of Neuroscience. 1996;16(14):4529–4535.

52. Bourgeois J-P, Jastreboff PJ, Rakic P. Synaptogenesis in visual cortex of normal and preterm monkeys: evidence for intrinsic regulation of synaptic overproduction. Proceedings of the National Academy of Sciences. 1989;86(11):4297–4301.

53. Brener ND, Billy JO, Grady WR. Assessment of factors affecting the validity of self-reported health-risk behavior among adolescents: evidence from the scientific literature. Journal of adolescent health. 2003;33(6):436–457.

54. Casey BJ, Cannonier T, Conley MI, et al. The Adolescent Brain Cognitive Development (ABCD) study: Imaging acquisition across 21 sites. Dev Cogn Neurosci. 08 2018;32:43–54. doi:10.1016/j.dcn.2018.03.001

55. Feldstein Ewing SW, Chang L, Cottler LB, Tapert SF, Dowling GJ, Brown SA. Approaching Retention within the ABCD Study. Dev Cogn Neurosci. Aug 2018;32:130–137. doi:10.1016/j.dcn.2017.11.004

56. Karcher NR, Barch DM. The ABCD study: understanding the development of risk for mental and physical health outcomes. Neuropsychopharmacology. 2021/01/01 2021;46(1):131–142. doi:10.1038/s41386-020-0736-6

57. Hagler Jr DJ, Hatton S, Cornejo MD, et al. Image processing and analysis methods for the Adolescent Brain Cognitive Development Study. Neuroimage. 2019;202:116091.

58. Uban KA, Horton MK, Jacobus J, et al. Biospecimens and the ABCD study: Rationale, methods of collection, measurement and early data. Dev Cogn Neurosci. Aug 2018;32:97–106. doi:10.1016/j.dcn.2018.03.005

59. Zucker RA, Gonzalez R, Feldstein Ewing SW, et al. Assessment of culture and environment in the Adolescent Brain and Cognitive Development Study: Rationale, description of measures, and early data. Dev Cogn Neurosci. Aug 2018;32:107–120. doi:10.1016/j.dcn.2018.03.004

60. Barch DM, Albaugh MD, Avenevoli S, et al. Demographic, physical and mental health assessments in the adolescent brain and cognitive development study: Rationale and description. Dev Cogn Neurosci. Aug 2018;32:55–66. doi:10.1016/j.dcn.2017.10.010

61. Luciana M, Bjork JM, Nagel BJ, et al. Adolescent neurocognitive development and impacts of substance use: Overview of the adolescent brain cognitive development (ABCD) baseline neurocognition battery. Dev Cogn Neurosci. Aug 2018;32:67–79. doi:10.1016/j.dcn.2018.02.006

62. Cornejo MD, Makowski C, Dick AS, et al. Image processing and analysis methods for the Adolescent Brain Cognitive Development Study. NeuroImage (Orlando, Fla). 2019;202:116091–116091. doi:10.1016/j.neuroimage.2019.116091

63. Fischl B, Salat DH, Busa E, et al. Whole brain segmentation: automated labeling of neuroanatomical structures in the human brain. Neuron. 2002;33(3):341–355.

64. Kind AJH, Jencks S, Brock J, et al. Neighborhood socioeconomic disadvantage and 30-day rehospitalization: a retrospective cohort study. Annals of internal medicine. 2014;161(11):765–774. doi:10.7326/M13-2946

65. Baurley JW, Edlund CK, Pardamean CI, Conti DV, Bergen AW. Smokescreen: a targeted genotyping array for addiction research. BMC genomics. 2016;17(128):145–145. doi:10.1186/s12864-016-2495-7

66. Corresponding GPC. An integrated map of genetic variation from 1,092 human genomes. Nature. 2012;491(7422):56–65.

67. Howie B, Donnelly P, Marchini J. 1,000 Genomes haplotypes—Phase 3 integrated variant set release in NCBI build 37 (hg19) coordinates. 2015.

68. Lam M, Awasthi S, Watson HJ, et al. RICOPILI: Rapid Imputation for COnsortias PIpeLIne. Bioinformatics. Feb 1 2020;36(3):930–933. doi:10.1093/bioinformatics/btz633

69. Nielsen TT, Duan J, Levey DF, et al. Disentangling the shared genetics of ADHD, cannabis use disorder and cannabis use and prediction of cannabis use disorder in ADHD. medRxiv. 2024:2024.02. 22.24303124.

70. Zhou H, Kember RL, Deak JD, et al. Multi-ancestry study of the genetics of problematic alcohol use in over 1 million individuals. Nature Medicine. 2023/12/01 2023;29(12):3184–3192. doi:10.1038/s41591-023-02653-5

71. Choi SW, O’Reilly PF. PRSice-2: Polygenic Risk Score software for biobank-scale data. Gigascience. 2019;8(7)doi:10.1093/gigascience/giz082

72. Lee JJ, Wedow R, Okbay A, et al. Gene discovery and polygenic prediction from a genome-wide association study of educational attainment in 1.1 million individuals. Nature genetics. 2018;50(8):1112–1121.

73. Turley P, Walters RK, Maghzian O, et al. Multi-trait analysis of genome-wide association summary statistics using MTAG. Nature genetics. 2018;50(2):229–237.

74. Brener ND, Billy JO, Grady WR. Assessment of factors affecting the validity of self-reported health-risk behavior among adolescents: evidence from the scientific literature. J Adolesc Health. Dec 2003;33(6):436–57. doi:10.1016/s1054-139x(03)00052-1

75. Riley AW. Evidence that school-age children can self-report on their health. Ambulatory Pediatrics. 2004;4(4):371–376.

76. Nagata JM, Cortez CA, Cattle CJ, et al. Screen Time Use Among US Adolescents During the COVID-19 Pandemic: Findings From the Adolescent Brain Cognitive Development (ABCD) Study. JAMA Pediatrics. 2022;176(1):94–96. doi:10.1001/jamapediatrics.2021.4334

77. Narvacan K, Treit S, Camicioli R, Martin W, Beaulieu C. Evolution of deep gray matter volume across the human lifespan. Hum Brain Mapp. Aug 2017;38(8):3771–3790. doi:10.1002/hbm.23604

78. Tiemeier H, Lenroot RK, Greenstein DK, Tran L, Pierson R, Giedd JN. Cerebellum development during childhood and adolescence: a longitudinal morphometric MRI study. Neuroimage. Jan 1 2010;49(1):63–70. doi:10.1016/j.neuroimage.2009.08.016

79. Wierenga L, Langen M, Ambrosino S, van Dijk S, Oranje B, Durston S. Typical development of basal ganglia, hippocampus, amygdala and cerebellum from age 7 to 24. NeuroImage. 2014/08/01/ 2014;96:67–72. 10.1016/j.neuroimage.2014.03.072

80. Tamnes CK, Herting MM, Goddings AL, et al. Development of the Cerebral Cortex across Adolescence: A Multisample Study of Inter-Related Longitudinal Changes in Cortical Volume, Surface Area, and Thickness. J Neurosci. Mar 22 2017;37(12):3402–3412. doi:10.1523/jneurosci.3302-16.2017

81. Burnham KP, Anderson DR. Multimodel inference: understanding AIC and BIC in model selection. Sociological methods & research. 2004;33(2):261–304.

82. Bates D, Mächler M, Bolker B, Walker S. Fitting linear mixed-effects models using lme4. arXiv preprint arXiv:14065823. 2014;

83. R Core Team R. R: A language and environment for statistical computing. 2013;

84. Benjamini Y, Hochberg Y. Controlling the false discovery rate: a practical and powerful approach to multiple testing. Journal of the Royal statistical society: series B (Methodological). 1995;57(1):289–300.

85. Sripada K, Bjuland KJ, Sølsnes AE, et al. Trajectories of brain development in school-age children born preterm with very low birth weight. Sci Rep. Oct 22 2018;8(1):15553. doi:10.1038/s41598-018-33530-8

86. Brumbaugh JE, Conrad AL, Lee JK, et al. Altered brain function, structure, and developmental trajectory in children born late preterm. Pediatr Res. Aug 2016;80(2):197–203. doi:10.1038/pr.2016.82

87. Nivins S, Kennedy E, McKinlay C, et al. Size at birth predicts later brain volumes. Scientific Reports. 2023/08/01 2023;13(1):12446. doi:10.1038/s41598-023-39663-9

88. Shaw P, Malek M, Watson B, Sharp W, Evans A, Greenstein D. Development of cortical surface area and gyrification in attention-deficit/hyperactivity disorder. Biological psychiatry. 2012;72(3):191–197.

89. Lüdecke D. ggeffects: Tidy data frames of marginal effects from regression models. Journal of Open Source Software. 2018;3(26):772.

