## Supplementary for "Long-Term Impact of Digital Media on Brain Development in Children"

**Tables**

**eTable 1**. AIC BIC values for linear and quadratic models to describe the relationship between age and brain regions

|  | **Models** | | | |
| --- | --- | --- | --- | --- |
| **Brain regions** | **Linear** | | **Quadratic** | |
|  | AIC | BIC | AIC | BIC |
| Total cortical surface area | 13538 | 13888 | 13356 | 13714 |
| Striatum | 16566 | 16916 | 16450 | 16808 |
| Cerebellum | 13809 | 14160 | 13069 | 13427 |

Abbreviations: AIC, Akaike Information Criterion; BIC, Bayesian Information Criterion. The model with lower AIC and BIC values was considered a better fit (at least by 10 points less than the other model).

**eTable 2** Sex-specific effect on brain development

| **Brain regions** | **Age** | | **Age^2^** | | **Age x sex** | | **Age^2^ x sex** | |
| --- | --- | --- | --- | --- | --- | --- | --- | --- |
|  | **β (SE)** | **P** | **β (SE)** | **p** | **β (SE)** | **P** | **β (SE)** | **P** |
| Global cortical surface area (mm^2^) | 0.39  (0.02) | < 0.001 | -0.02  (0.001) | < 0.001 | -0.21  (0.03) | < 0.001 | -0.01  (0.001) | < 0.001 |
| Striatum volume (mm^3^) | 0.07  (0.03) | 0.009 | -0.004  (0.001) | 0.002 | -0.05  (0.03) | 0.11 | 0.002  (0.001) | 0.14 |
| Cerebellum volume (mm^3^) | 0.22  (0.02) | < 0.001 | -0.01  (0.001) | < 0.001 | -0.36  (0.03) | < 0.001 | 0.02  (0.001) | < 0.001 |

P values presented are uncorrected for multiple comparisons.

**eTable 3** Absolute brain sizes at different timepoints in boys and girls

| **Brain regions** | **Boys** | **Girls** |
| --- | --- | --- |
|  | **mean (SD)** | **mean (SD)** |
| Global cortical surface area (mm^2^)  T_0_  T_2_  T_4_ | 198453 (16690)  198779 (16745)  198617 (16966) | 181951 (15266)  181829 (15234)  179961 (14969) |
| Cerebellum volume (mm^3^)  T_0_  T_2_  T_4_ | 152279 (13319)  154307 (12210)  156495 (16581) | 140407 (11993)  142075 (12210)  141433 (12286) |

**eTable 4** Association between sex-specific individual digital media exposure and brain outcomes in children aged 9-11 years with 4 years of follow-ups.

| **Brain regions** | **Social media usage** | | **Time**  **(linear)** | | **Time**  **(Quadrant)** | | **Social media usage x Time x Sex** | | **Social media usage x Time^2^ x Sex** | |
| --- | --- | --- | --- | --- | --- | --- | --- | --- | --- | --- |
|  | **β (SE)** | **p** | **β (SE)** | **P** | **β (SE)** | **P** | **β (SE)** | **P** | **β (SE)** | **P** |
| Global cortical surface area (mm^2^) | 0.001  (0.02) | 0.91 | 0.08  (0.02) | < 0.001 | -0.07  (0.01) | <0.001 | 0.03  (0.01) | 0.006 | -0.02  (0.006) | 0.01 |
| Striatum  Volume (mm^3^) | -0.001  (0.02) | 0.94 | -0.01  (0.01) | 0.24 | -0.005  (0.01) | 0.69 | -0.004  (0.01) | 0.70 | 0.003  (0.01) | 0.62 |
| Cerebellum  Volume (mm^3^) | 0.02  (0.01) | 0.05 | 0.15  (0.01) | <0.001 | -0.04  (0.008) | <0.001 | 0.03  (0.009) | 0.001 | -0.01  (0.006) | 0.08 |
|  | **Playing video games** | | **Time**  **(linear)** | | **Time**  **(Quadrant)** | | **Playing video games x Time x Sex** | | **Playing video games x Time^2^ x Sex** | |
|  | **β (SE)** | **p** | **β (SE)** | **P** | **β (SE)** | **P** | **β (SE)** | **P** |  |  |
| Global cortical surface area (mm^2^) | -0.04  (0.02) | <0.001 | 0.04  (0.01) | 0.001 | -0.06  (0.001) | <0.001 | 0.01  (0.01) | 0.42 | -0.001  (0.006) | 0.77 |
| Striatum  Volume (mm^3^) | -0.008  (0.02) | 0.59 | 0.003  (0.01) | 0.80 | -0.007  (0.007) | 0.33 | -0.004  (0.01) | 0.72 | 0.007  (0.007) | 0.32 |
| Cerebellum  Volume (mm^3^) | 0.004  (0.01) | 0.74 | 0.13  (0.01) | <0.001 | -0.03  (0.006) | <0.001 | -0.004  (0.01) | 0.96 | 0.006  (0.006) | 0.32 |
|  | **Watching television** | | **Time**  **(linear)** | | **Time**  **(Quadrant)** | | **Watching television x Time x Sex** | | **Watching television x Time^2^ x Sex** | |
|  | **β (SE)** | **p** | **β (SE)** | **p** | **β (SE)** | **P** | **β (SE)** | **P** | **β (SE)** | **P** |
| Global cortical surface area (mm^2^) | -0.03  (0.01) | 0.01 | 0.05  (0.02) | 0.005 | -0.06  (0.009) | <0.001 | 0.002  (0.008) | 0.50 | 0.001  (0.004) | 0.72 |
| Striatum  Volume (mm^3^) | -0.01  (0.01) | 0.33 | -0.02  (0.02) | 0.25 | 0.006  (0.01) | 0.53 | -0.004  (0.009) | 0.63 | 0.007  (0.005) | 0.19 |
| Cerebellum  Volume (mm^3^) | 0.01  (0.01) | 0.31 | 0.11  (0.01) | <0.001 | -0.02  (0.008) | 0.03 | 0.001  (0.007) | 0.79 | 0.004  (0.004) | 0.28 |

P values presented are uncorrected for multiple comparisons.

**eTable 5** Association between average social media/video gaming exposure of first two time points (T_0_ and T_1_) and changes in cerebellum volumes between T_2_ and T_4_ in the overall cohort

| **Brain regions** | **Social media usage at T_avg01_** | | **Time** | | **SES** | | **cogPGS** | | **Social media usage at T_avg01_ x Time** | |
| --- | --- | --- | --- | --- | --- | --- | --- | --- | --- | --- |
|  | **β (SE)** | **P** | **β (SE)** | **P** | **β (SE)** | **P** | **β (SE)** | **P** | **β (SE)** | **P** |
| Cerebellum  Volume (mm^3^) | 0.02  (0.01) | 0.07 | 0.09  (0.01) | < 0.001 | 0.03  (0.02) | 0.08 | -0.02  (0.01) | 0.30 | -0.01  (0.004) | 0.10 |
|  | **Playing video games at T_avg01_** | | **Time** | | **SES** | | **cogPGS** | | **Playing video games at T_avg01_ x Time** | |
|  | **β (SE)** | **P** | **β (SE)** | **P** | **β (SE)** | **P** | **β (SE)** | **P** | **β (SE)** | **P** |
| Cerebellum  Volume (mm^3^) | -0.02  (0.01) | 0.14 | 0.06  (0.01) | < 0.001 | 0.04  (0.02) | 0.07 | -0.03  (0.02) | 0.09 | 0.02  (0.01) | < 0.001 |

Abbreviations: SES, socioeconomic status; cogPGS, polygenic scores for cognitive performance. T_avg01_ = average of social media/playing video games atT_0_ and T_1_

P values presented are uncorrected for multiple comparisons.

**eTable 6** Association between social media usage at T_0_ and total change in cerebellum volumes (delta) adjusting for covariates

| **Brain regions** | **Social media usage at T_0_** | | **SES** | | **PGS** | |
| --- | --- | --- | --- | --- | --- | --- |
|  | **β (SE)** | **p** | **β (SE)** | **P** | **β (SE)** | **P** |
| Cerebellum (T_4_ – T_0_) (mm^3^) | -0.03  (0.02) | 0.16 | 0.06  (0.03) | 0.02 | 0.04  (0.02) | 0.08 |

Model: Delta (T_4_ – T_0_) ~ social media at T_0_ + age + SES + PGS + 20PCs + Total brain volumes at T_0_ + (1| sites)

Abbreviations: SES, socioeconomic status; cogPGS, polygenic scores for cognitive performance.

**eTable 7** Association between social media/video gaming exposure and cerebellum volumes in children aged 9-11 years with 4 years of follow-ups in the overall cohort excluding children born preterm or with ADHD diagnosis

| **Brain regions** | **Social media usage** | | **Time**  **(linear)** | | **Time**  **(Quadrant)** | | **SES** | | **cogPGS** | | **Social media usage x Time** | | **Social media usage x Time^2^** | |
| --- | --- | --- | --- | --- | --- | --- | --- | --- | --- | --- | --- | --- | --- | --- |
|  | **β (SE)** | **P** | **β (SE)** | **P** | **β (SE)** | **p** | **β (SE)** | **P** | **β (SE)** | **P** | **β (SE)** | **P** | **β (SE)** | **P** |
| Cerebellum  Volume (mm^3^) | 0.03  (0.01) | 0.04 | 0.07  (0.009) | <0.001 | 0.02  (0.006) | 0.003 | 0.04  (0.02) | 0.07 | -0.01  (0.02) | 0.47 | 0.02  (0.003) | <0.001 | -0.02  (0.004) | <0.001 |
|  | **Playing video games** | | **Time**  **(linear)** | | **Time**  **(Quadrant)** | | **SES** | | **cogPGS** | | **Playing video games x Time** | | **Playing video games x Time^2^** | |
|  | **β (SE)** | **P** | **β (SE)** | **P** | **β (SE)** | **P** | **β (SE)** | **P** | **β (SE)** | **P** | **β (SE)** | **P** |  |  |
| Cerebellum  Volume (mm^3^) | 0.002  (0.01) | 0.86 | 0.11  (0.009) | <0.001 | -0.02  (0.006) | <0.001 | 0.06  (0.02) | 0.006 | -0.004  (0.02) | 0.81 | -0.01  (0.01) | 0.13 | 0.01  (0.003) | 0.001 |

P values presented are uncorrected for multiple comparisons. Abbreviations: SES, socioeconomic status; cogPGS, polygenic scores for cognitive performance.

**eTable 8** Association between social media/video gaming exposure and cerebellum volumes in children aged 9-11 years with 4 years of follow-ups in children with MRI data available across all three visits (n=1462)

| **Brain regions** | **Social media usage** | | **Time**  **(linear)** | | **Time**  **(Quadrant)** | | **SES** | | **cogPGS** | | **Social media usage x Time** | | **Social media usage x Time^2^** | |
| --- | --- | --- | --- | --- | --- | --- | --- | --- | --- | --- | --- | --- | --- | --- |
|  | **β (SE)** | **P** | **β (SE)** | **P** | **β (SE)** | **p** | **β (SE)** | **P** | **β (SE)** | **P** | **β (SE)** | **P** | **β (SE)** | **P** |
| Cerebellum  Volume (mm^3^) | 0.03  (0.02) | 0.16 | 0.08  (0.01) | <0.001 | 0.01  (0.006) | 0.02 | 0.02  (0.04) | 0.62 | -0.03  (0.03) | 0.42 | 0.01  (0.01) | 0.09 | -0.01  (0.004) | 0.002 |
|  | **Playing video games** | | **Time**  **(linear)** | | **Time**  **(Quadrant)** | | **SES** | | **cogPGS** | | **Playing video games x Time** | | **Playing video games x Time^2^** | |
|  | **β (SE)** | **P** | **β (SE)** | **P** | **β (SE)** | **P** | **β (SE)** | **P** | **β (SE)** | **P** | **β (SE)** | **P** |  |  |
| Cerebellum  Volume (mm^3^) | -0.02  (0.02) | 0.34 | 0.13  (0.01) | <0.001 | -0.03  (0.007) | <0.001 | 0.06  (0.04) | 0.12 | -0.06  (0.03) | 0.10 | -0.02  (0.007) | 0.001 | 0.02  (0.004) | <0.001 |

P values presented are uncorrected for multiple comparisons. Abbreviations: SES, socioeconomic status; cogPGS, polygenic scores for cognitive performance.

**Figures**

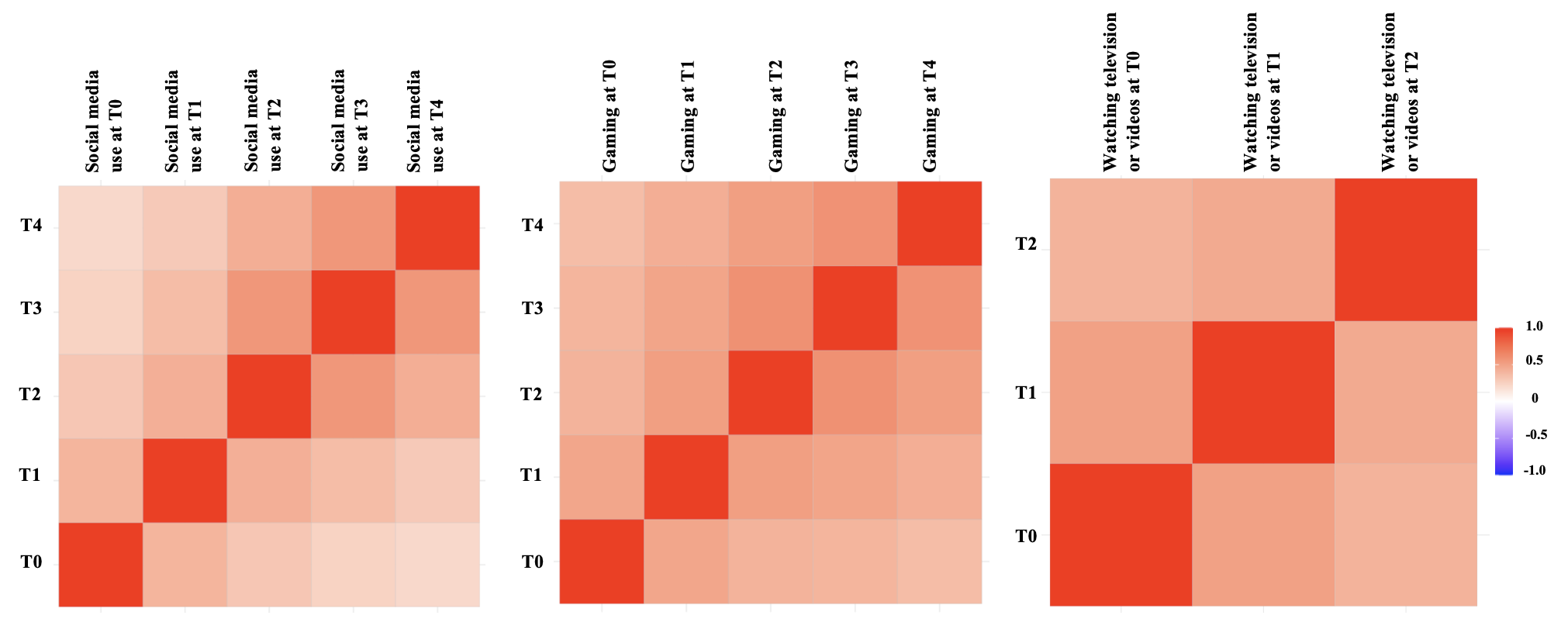

**eFigure 1** Correlation between individual digital media (i.e., social media usage, playing video games, or watching television/videos) usage across four different waves of follow-up
